# Organizational impact of an ID NOW™ COVID-19 point-of-care testing for SARS-CoV2 detection in a maternity ward

**DOI:** 10.1101/2022.08.29.22279161

**Authors:** Jean-Claude Nguyen Van, Benoît Pilmis, Amir Khaterchi, Olivier Billuart, Gauthier Péan De Ponfilly, Alban Le Monnier, Elie Azria, Assaf Mizrahi

## Abstract

**Background:** SARS-CoV-2 has been responsible for more than 550 million cases of COVID-19 worldwide. RT-PCR is considered the “gold standard” for the diagnosis of patients suspected of having COVID-19. During the heightened waves of the pandemic, more rapid tests have been required. Point-of-care tests (POCT) for COVID-19 include antigen tests, serological tests, and other molecular-based platforms. The ID NOW™ COVID-19 assay (Abbott) performs an isothermal gene amplification of a target encoding the RNA-dependent RNA polymerase of SARSCoV-2. The main objective of this study was to evaluate the organizational impact following the implementation of a POC testing platform ID NOW™ in a maternity ward.

**Materials and Methods:** This retrospective study included pregnant women admitted for Groupe Hospitalier Paris Saint-Joseph Paris. The study was conducted over 2 periods lasting 6 months each. The first period (P1) corresponded to the 2nd wave in France (July to December 2020) whereas the second (P2) period focused on the 3rd wave (February to July 2021). During P1, viral detection was performed by RT-PCR at the hospital’s laboratory. During P2, it was performed with the ID NOW™ COVID-19 test directly in the delivery room by nursing staff after training and certification. Our primary endpoint was the length of time in the birth room from admission to discharge in the postpartum period.

**Results:** 2447 pregnant women were included, 1053 during P1 and 1394 during P2. The median age, percentage of singleton pregnancies, mean gestational age, percentage of nulliparous individuals, percentage of vaginal deliveries, and COVID19 positivity rate were comparable between the two periods. During P2, the length of stay in the delivery room was significantly shorter than during P1 (17.9 *vs* 14.7 hours, *p*<0.001).

**Conclusion:** Analysis of the data from this study following the implementation of the ID NOW™ POCT in the maternity ward indicates a significant decrease in the length of stay in the birth room. This outcome needs to be confirmed in a multicenter cohort, in particular to precise the specific impact of COVID-19 care on delays.

## Introduction

SARS-CoV-2 has been responsible for more than 550 million cases of COVID-19 worldwide. The onset of this COVID-19 pandemic revealed our limited supply of molecular tests. Increasing COVID-19 testing was identified as an essential component of the pandemic control strategy. Thus, rapid and accurate tests were urgently needed. Therefore, molecular testing in point-of-care strategy in maternity ward became apparent. Indeed, point-of care testing is associated with large reductions in time to results and could lead to improvements the work flow in birth room.

The implementation of the POCT for influenza for several years and coronavirus in the emergency department of our hospital has allowed us to benefit from this significant experience ^1,2^. The ID NOW COVID-19 test is a rapid molecular biology test based on isothermal gene amplification reaction (NEAR). Pregnant women, particularly those with associated comorbidities, seem to be at higher risk of severe complications of SARS-CoV-2 infection compared with non-pregnant women control ^3,4^.

RT-PCR is considered the “gold standard” for the diagnosis of patients suspected of having COVID-19. During the heightened waves of the pandemic, more rapid tests have been required. Point-of-care tests (POCT) for COVID-19 include antigen tests, serological tests, and other molecular-based platforms. The development of rapid molecular diagnostic tests that have sensitivity and specificity comparable to the current gold standard techniques can significantly aid testing expansion ^5,6^. Thus, the ID NOW™ COVID-19 assay (Abbott) performs a rapid isothermal gene amplification of a target encoding the RNA-dependent RNA polymerase of SARSCoV-2. In a previous study, we have shown that this ID NOW™ assay has comparable performance to RT-PCR ^7^. The main objective of this study was to evaluate the organizational impact following the implementation of a POC testing platform ID NOW™ in our maternity ward.

## Materials and Methods

### Study design and setting

This was a retrospective study in a single center. All parturients were recruited from the maternity ward of a tertiary hospital called Groupe Hospitalier Paris Saint-Joseph, Paris, France.

This retrospective study included pregnant women admitted for childbirth in our hospital in Paris. The study was conducted over 2 periods lasting 6 months each. The first period (P1) corresponded to the 2nd wave (July to December 2020) whereas the second (P2) period focused on the 3rd wave (February to July 2021). During P1, viral detection was performed by RT-PCR at the hospital’s laboratory. During P2, viral detection was performed with the ID NOW™ COVID-19 test directly in the delivery room by nursing staff after training and certification. The assay was performed directly from the dry swab obtained from pregnant women at the beginning of delivery.

The primary endpoint was the length of time in the birth room from admission to discharge in the postpartum period The second endpoint was the percentage of COVID tests obtained after the time from onset of labor to delivery added to 2 hours of legal delay (2h *post-partum*).

### Testing methods

The ID NOW COVID-19 assay (Abbott, Chicago, Il, USA) is an isothermal nucleic acid amplification-based. This assay is a rapid molecular diagnostic test which uses nicking enzyme amplification reaction (NEAR) technology for the detection of SARS-CoV-2 RNA, targeting the RdRp gene ^8^. Samples can only be tested one at a time. Thus, nasopharyngeal swabs were collected with a flexible nasopharyngeal flocked swabs and during period 2, swabs were directly tested on the ID NOW™ COVID-19 assay at POC by ED trained nurses or midwifes previously trained and certified for use it. Following an initial 3 min warm-up of the test system, a dry swab is added to elution buffer in the sample receiver and then mixed for 10 s. Using the sample transfer device, 200 μL of sample is transferred into the test cartridge, the lid is closed, and the instrument automatically initializes the assay, which runs for 10 min. The ID NOW™ test provides a qualitative result and does not report cycle threshold (Ct) values to the user.

The instrument software interprets amplification data and final results are reported as positive, negative, or invalid. Samples that yield an initial invalid result are repeated. If an invalid result is generated twice, the final result is reported as invalid.

Connectivity between POCT instruments and the electronic medical record is provided by the middleware AegisPOC™ Point of Care Management Solutions.

### Statistical analysis

The percentages were calculated based on documented data (missing data were excluded from the percentages). Inter-group comparisons were made using the Mann-Whitney test for quantitative variables and the Fisher exact test for qualitative variables. Statistical analysis was done with StatView software (version 5.0). All tests were two-tailed and p values less than 0.05 (calculated by **c** test, Student’s *t* test, or Mann-Whitney test) were considered significant.

### Practitioners’ perceptions regarding COVID-19 POC implementation in the Maternity wards

User satisfaction with the ID NOW™ COVID-19 test was assessed through a qualitative/quantitative questionnaire that was completed by participating nurses and midwifes

### Ethical statement

This study followed the guidelines of the Standards for Reporting of Diagnostic Accuracy studies (STARD) and was previously approved by the clinical ethics committee of Saint-Joseph hospital called “Groupe Ethique Recherche Medicale” which issued a favorable ethical opinion and registered in the single protocol DELOCOVIDMATER NCT05175716 of clinical trial.gov (https://clinicaltrials.gov/ct2/show/NCT05175716). Informed oral consent for participation was obtained from each participant, in accordance with French law.

## Results

2447 pregnant women were included, 1053 during P1 and 1394 during P2. Among the exclusion criteria were, scheduled deliveries, prelabor caesarean sections, parturients with missing data and parturients with preterm birth (<37 weeks gestation) were excluded. Thus, 1066 parturients were included, 449 during P1 and 617 during P2 (Figure 1).

**Figure 1:**
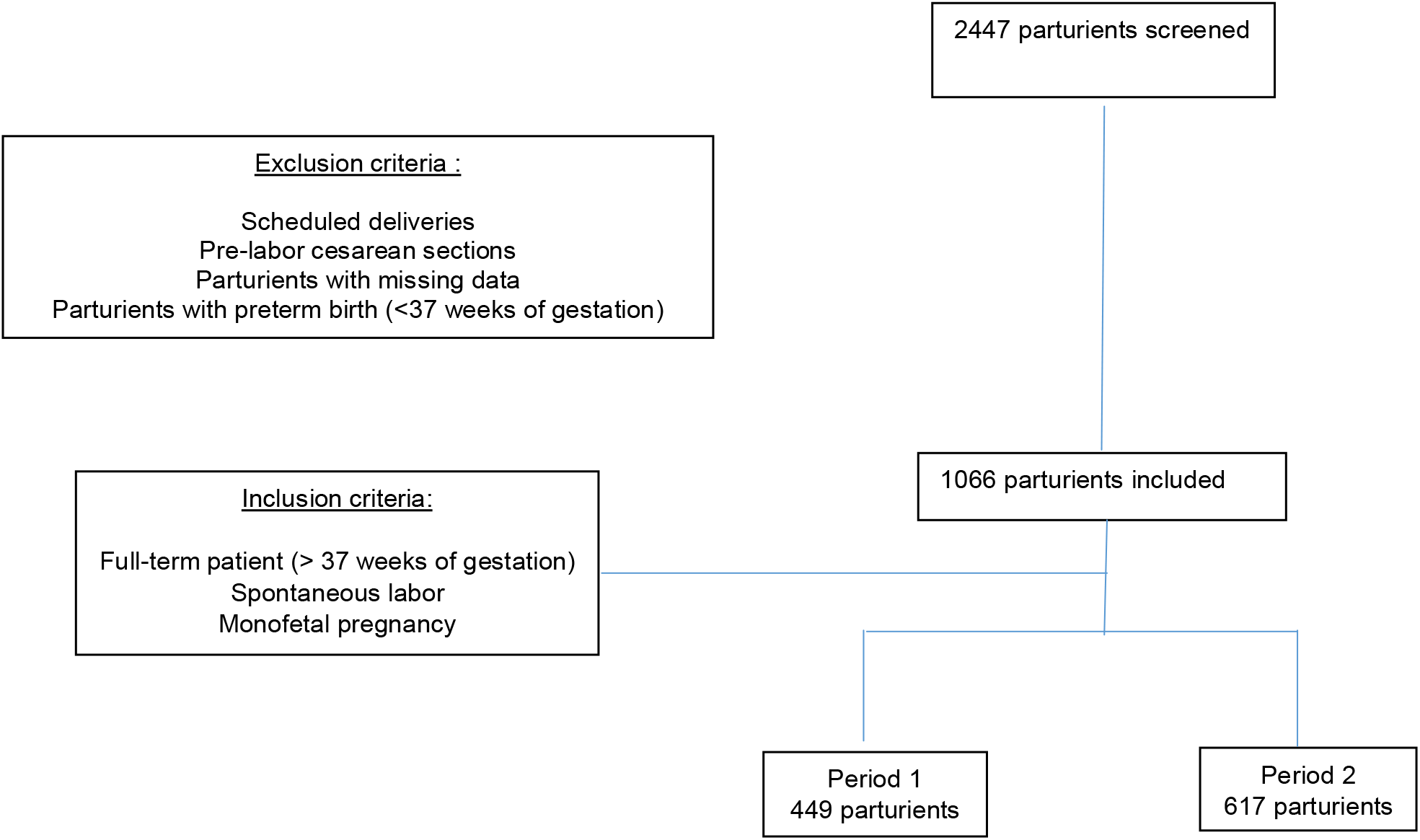
Patient flow chart.

The median age, percentage of singleton pregnancies, mean gestational age, percentage of nulliparous individuals, percentage of vaginal deliveries, and COVID19 positivity rate were comparable between the two periods (table 1). During P2, the median time in birthroom was significantly shorter than during P1 (10.40 vs 14.59 hours, p<0.001). For the nulliparous women, the median time in birth room was always significantly shorter than during P1 (16.08 vs 19.02 hours, p<0.001).

**Table 1.**
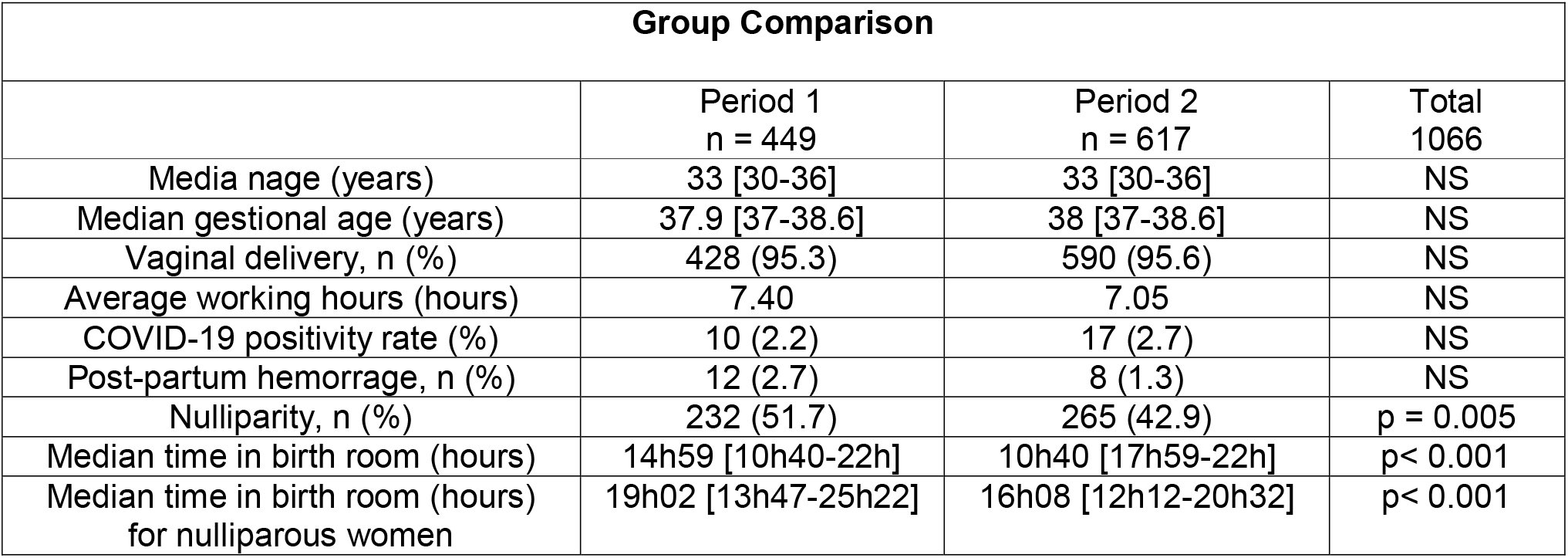
Comparison of groups during period 1 and period 2

The percentages of COVID tests obtained after the time from onset of labor to delivery added to 2 hours of medical *post-partum* were 73/448 (19.4%) and 4/596 (0.6%) for period 1 and period 2, respectively (p <0.05).

At the end of the study, users’ opinions on the COVID ID NOW test were collected. Survey responses are shown in Table 2. A total of sixty-three practitioners participated to the survey including forty-four midwifes (63%), eighteen nurses (33%), one physician. The majority of practitioner*s* reported the implementation of ID NOW™COVID-19 POCT was straightforward. In addition, ease of use and duration of care scored high (90.6% and 93.0% respectively). Improved hygiene compliance and reduced fear of being contaminated scored well (75%) and the overall level of satisfaction with the use of the point-of-care was eighty-five percent (85%). POCT improved hygiene compliance and screened parturients as they entered the delivery room.

**Table 2.**
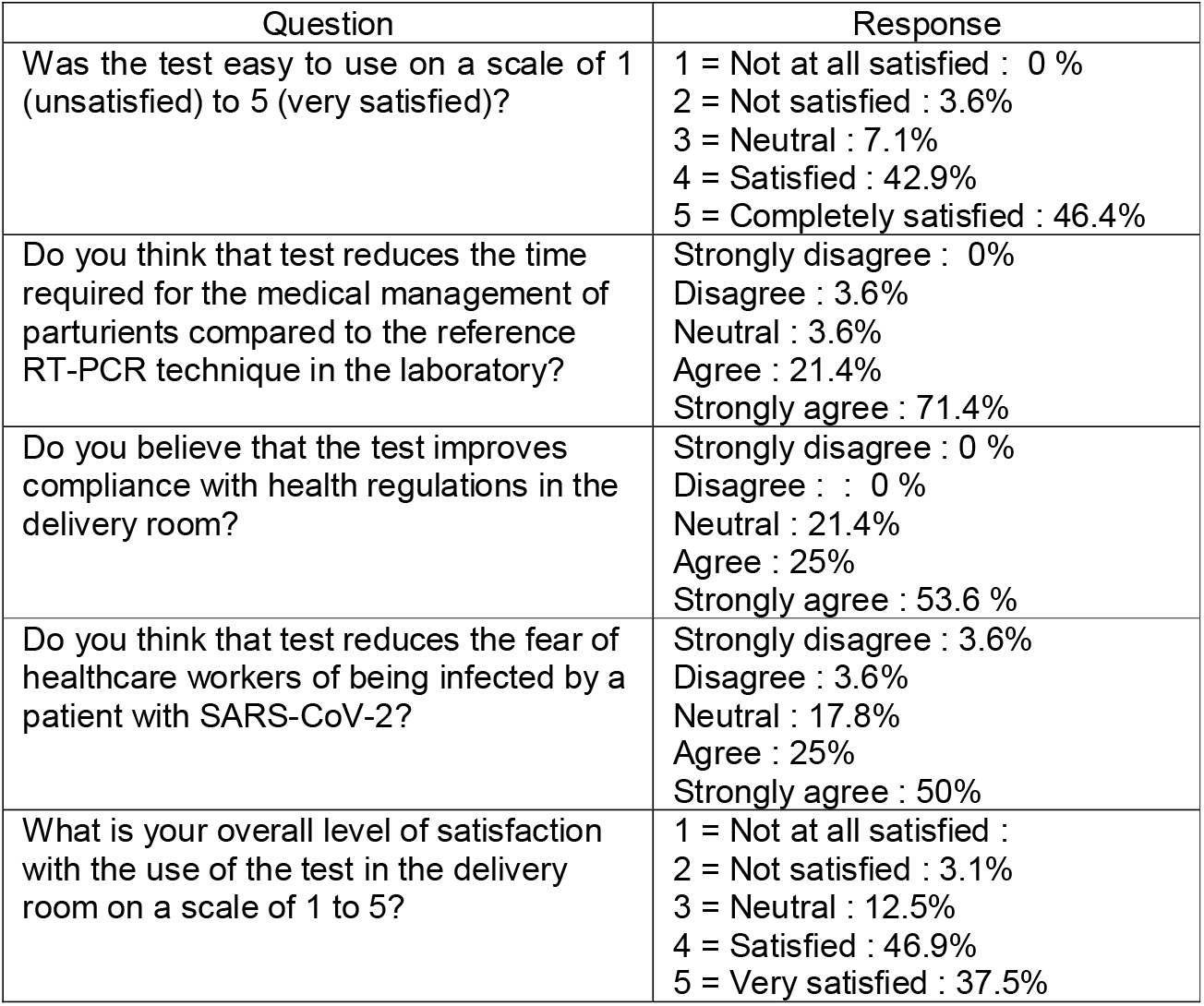
Evaluation of satisfaction and perception regarding COVID-19 POC implementation in the delivery room

| Question | Response |
| --- | --- |
| Was the test easy to use on a scale of 1 (unsatisfied) to 5 (very satisfied)? | 1 = Not at all satisfied : 0 %<br>2 = Not satisfied : 3.6%<br>3 = Neutral : 7.1%<br>4 = Satisfied : 42.9%<br>5 = Completely satisfied : 46.4% |
| Do you think that test reduces the time required for the medical management of parturients compared to the reference RT-PCR technique in the laboratory? | Strongly disagree : 0%<br>Disagree : 3.6%<br>Neutral : 3.6%<br>Agree : 21.4%<br>Strongly agree : 71.4% |
| Do you believe that the test improves compliance with health regulations in the delivery room? | Strongly disagree : 0 %<br>Disagree : : 0 %<br>Neutral : 21.4%<br>Agree : 25%<br>Strongly agree : 53.6 % |
| Do you think that test reduces the fear of healthcare workers of being infected by a patient with SARS-CoV-2? | Strongly disagree : 3.6%<br>Disagree : 3.6%<br>Neutral : 17.8%<br>Agree : 25%<br>Strongly agree : 50% |
| What is your overall level of satisfaction with the use of the test in the delivery room on a scale of 1 to 5? | 1 = Not at all satisfied :<br>2 = Not satisfied : 3.1%<br>3 = Neutral : 12.5%<br>4 = Satisfied : 46.9%<br>5 = Very satisfied : 37.5% |

## Discussion

Rapid and accurate detection of SARS-CoV-2 is essential to ensure early and appropriate parturient management. New diagnostic technologies with rapid turnaround and good performance are an opportunity to improve the organization in the birth room.

At the time of the first wave, the maternity care team was pressuring the laboratory to obtain COVID test results quickly. This pressure was even greater at night because the tests were only performed in central laboratory during the day. This led to disorganization in the laboratory and tension between the laboratory and the maternity ward. Therefore, we decided to implement ID NOW™ COVID-19 point of care testing for SARS-CoV2 detection in the maternity ward, using our point-of-care testing experience gained in the emergency department^1 2^.

The implementation of an automated system in point-of-care must follow rules and must not be done in a hurry. The nursing staff must be trained and authorized by medical referents of the laboratory. A project manager should be clearly named to monitor drift on a daily basis and communicate on positivity rates.

Implementation of ID NOW had led to a significantly lower time to result (TTR) compared to the classic procedure. The organizational impact following the implementation of a POC testing ID NOW™ platform for parturients could be evaluated by comparing the median time in the birth room with and without the ID NOW™ platform device. In order to obtain a homogeneous population the inclusion criteria were: full-term patient >37 weeks of gestation, spontaneous labor and mono-fetal pregnancy

We obtained a significant reduction in average time in the delivery room from 14.59 to 10.4 hours. This reduction of the length of time spent in the delivery room could be explained by the reduction of TTR. Indeed, in our settings, pregnant women had been sampled for COVID testing as soon as they went into labor. Once they have given birth, after a legal monitoring period of 2 hours, they are transferred to the postnatal suite.

During the first period of our study, 73 parturients out of 448 (19.4%) could not be transferred from the delivery room to the postnatal suite because they did not have their PCR results. Implementation of ID NOW allowed to decrease significantly this number to 4 parturients out of 596 (0.6%) during period 2. Nurses and midwifes who used the ID NOW™ COVID-19 assay received training from two experts, and were required to undertake a proficiency test prior to participation. Testing was completed in accordance with the manufacturers instructions. As reported by Oliver *et al*. the particularly crucial areas for POCT accreditation are method performance verification, internal and external quality assurance, staff training and competency, and continuous improvement ^9^. To this end, we have organized the supervision and support of the maternity ward staff with the help of a referral microbiologist, a quality assurance engineer and two referral technicians. This organization allowed us to ensure staff training, equipment maintenance, connectivity to the laboratory information system, quality control and external quality assurance procedures, all of which are required for accreditation according to the ISO 22870 standard ^10 11^

For connectivity, we used the AegisPOC™ Point of Care Management Solutions which is a web-based, open platform connecting point-of-care (POC) device. It allowed the laboratory to manage training certification of individuals performing POCT and share data from POC device on one middleware.

POCT is usually more expensive than testing performed in the central laboratory and requires a significant amount of support from the laboratory to ensure the quality testing and meet accreditation requirements ^11^. This point is important and requires careful consideration before embarking on a process that requires time, human resources (reference technicians, quality specialists and microbiologists) and material resources (connected prescription, middleware). It may be worthwhile to evaluate the cost-effectiveness in a future study and to assess the impact of the implementation of the COVID-19 POCT on direct and indirect hospital costs during the pandemic.

### Limitations

To our knowledge, this is the first study to evaluate the organizational impact in the delivery room but this work has several limitations. Firstly, it is a retrospective monocentric study. Results may be specific to the population of our hospital. Use of rapid diagnostic test at triage in other care settings would depend on the organization of each Maternity ward and, importantly, on the availability of compliant and trained staff. Secondly, the optimal assessment of the impact of COVID-19 POCT implementation should be evaluated in an unbiased randomized-controlled trial. Nevertheless, in the tense situation and in the expectation of the clinicians (gyneco-obstetricians), it was almost impossible to carry out a randomized study.

We observed excellent compliance by nurses and midwifes throughout the study, and the low number of invalid tests (< 4%) attests to good compliance with the procedure.

This is a collaborative and exemplary study. When we interviewed the medical and paramedical staff in the birthing suite, “no one would want to go back”. Additional studies are needed to confirm these results.

## Conclusion

Analysis of the data from this study following the implementation of the ID NOW™ POCT in the maternity ward indicates a significant decrease in the length of stay in the birth room. This outcome needs to be confirmed in a multicenter cohort, in particular to precise the specific impact of COVID-19 care on delays.

## Data Availability

All data produced in the present work are contained in the manuscript

## Why was this study conducted ?

This study aimed to improve organizational impact following the implementation of a point-of-care testing platform ID NOW™ in a maternity ward.

## Key findings

Implementation of ID NOW™ POCT had led to a significantly lower time to result (TTR) compared to the classic procedure. The organizational impact following the implementation of a POC testing ID NOW™ platform for parturients could be evaluated by comparing the median time that was significantly shorter with *versus* without the ID NOW™ platform device (10.40 vs 14.59 hours, p<0.001).

Based on the users’ opinion, the overall satisfaction level is high (85%).

## What does this study add to what is known ?

To our knowledge, this is the first study to evaluate the organizational impact following the implementation of a point-of-care testing platform ID NOW™ in the delivery room. This study showed a significant decrease in the length of stay in the birth room.

## Credit author statement

AM contributed to data curation, formal analysis, methodology, validation, writing original draft;

AK contributed to the curation, formal analysis,

BP contributed to the curation, formal analysis, methodology, validation;

OB data curation, formal analysis

GPP contributed to writing original draft.

ALM contributed to supervision, project administration.

JCNV contributed to conceptualization, data curation, formal analysis, methodology, project administration, supervision, validation, visualization, writing original draft;

JCNV drafted the manuscript to which all authors provided critical comments and a final consent to the publishing.

## References

1. Trabattoni E, Le V, Pilmis B, et al. Implementation of Alere i Influenza A & B point of care test for the diagnosis of influenza in an emergency department. Am J Emerg Med. Published online October 18, 2017. doi:10.1016/j.ajem.2017.10.046

2. Gerlier C, Pilmis B, Ganansia O, Le Monnier A, Nguyen Van JC. Clinical and operational impact of rapid point-of-care SARS-CoV-2 detection in an emergency department. Am J Emerg Med. 2021;50:713–718. doi:10.1016/j.ajem.2021.09.062

3. DeBolt CA, Bianco A, Limaye MA, et al. Pregnant women with severe or critical coronavirus disease 2019 have increased composite morbidity compared with nonpregnant matched controls. Am J Obstet Gynecol. 2021;224(5):510.e1-510.e12. doi:10.1016/j.ajog.2020.11.022

4. Jamieson DJ, Rasmussen SA. An update on COVID-19 and pregnancy. Am J Obstet Gynecol. 2022;226(2):177–186. doi:10.1016/j.ajog.2021.08.054

5. Valera E, Jankelow A, Lim J, et al. COVID-19 Point-of-Care Diagnostics: Present and Future. ACS Nano. 2021;15(5):7899–7906. doi:10.1021/acsnano.1c02981

6. Vandenberg O, Martiny D, Rochas O, van Belkum A, Kozlakidis Z. Considerations for diagnostic COVID-19 tests. Nat Rev Microbiol. 2021;19(3):171–183. doi:10.1038/s41579-020-00461-z

7. Nguyen Van JC, Gerlier C, Pilmis B, et al. Prospective Evaluation of ID NOW COVID-19 Assay Used as Point-of-Care Test in an Emergency Department. Emergency Medicine; 2021. doi:10.1101/2021.03.29.21253909

8. Abbott Laboratories. ID Now COVID-19 package insert. Abbott Labora-tories, Chicago, IL.https://www.fda.gov/media/136525/download.

9. Oliver P, Fernandez-Calle P, Buno A. POCT Accreditation ISO 15189 and ISO 22870: Making the Point. EJIFCC. 2021;32(2):131–139.

10. Florkowski C, Don-Wauchope A, Gimenez N, Rodriguez-Capote K, Wils J, Zemlin A. Point-of-care testing (POCT) and evidence-based laboratory medicine (EBLM) – does it leverage any advantage in clinical decision making? Crit Rev Clin Lab Sci. 2017;54(7-8):471–494. doi:10.1080/10408363.2017.1399336

11. Shaw JLV. Practical challenges related to point of care testing. Pract Lab Med. 2016;4:22–29. doi:10.1016/j.plabm.2015.12.002

